# Analysing air particle quantity in a dental primary care setting

**DOI:** 10.1101/2020.08.12.20173450

**Authors:** A. J. Bates, D. R. Bates

## Abstract

**Objectives:** This study was undertaken to assess the amount of dental aerosol created in a primary care dental surgery.

**Methods:** Two particle meters were placed a set distances round a volunteer patient whilst undergoing simulated dental treatment using a high speed dental handpiece, and 3-in-1 air/water syringe, moisture control was managed with high volume suction and a saliva ejector. Measurement were taken every thirty seconds with the surgery environment set a neutral ventilation and with the windows open plus fan assistance.

**Results:** From the cessation of aerosol generation it took between 6 and 19 minutes for the surgery to return to baseline. The ventilated surgery had faster aerosol dispersal, returning to background levels within 5 minutes.

**Conclusion:** It is concluded for the surgery under investigation the dental aerosol had dissipated after 30 minutes using HVS and optimal surgery.

## Introduction

Dental surgeries have previously been investigated (1-6) for the levels of contamination both directly and indirectly from dental created aerosols. Most of these papers have concentrated upon bacterial contamination, with little on viral contamination. These papers have influenced the development of the current (pre Severe Acute Respiratory Syndrome Coronavirus-2 (COVID-19)) cross infection control standard operating procedures (SOPs) (7, 8) undertaken in dental practice and detailed in HTM 01-05(9). A particular problem for clinicians now being the recommended 1-hour fallow time following a dental AGP to allow the surgery to clear of any airborne virus.

With the onset of the 2020 COVID19 worldwide pandemic there has an increase in the research on the transmission of respiratory virus in healthcare settings, with almost all being in secondary hospital settings. These has been little investigation of primary care dental clinics. This is particularly important as respiratory transmission has been cited as a major source of COVID19 infection and number of medical Aerosol Generating Procedures (m-AGPs) have been named as being high risk by the WHO (10, 11). In the UK, dentistry has been included in list of aerosol creating procedures that may transmit respiratory virus.

Unfortunately, there is little information on the level of dental created aerosols and the risk they create. This is important as a dental aerosol is clean water that is contaminated by contact with a patient’s mucosal, oral, and nasal secretions. This is quite different from m-AGPs which are almost pure mucosal secretions. It is not in dispute that m-AGPs carry risk of virus transfer but is far from clear if dental Aerosol Generating Procedures (d-AGP) carry the same level of risk.

In the current COVID19 pandemic van Doremalen and coworkers (12) paper on the length of time that an aerosol can maintain viability COVID19 virus is quoted as being a median half-life on stainless steel and plastic of 5.6 and 6.8 hours respectively. However, this is under ideal experimental conditions using a Goldberg drum constantly agitating the aerosol whilst maintaining high temperature and 100% humidity. Real world conditions can reasonably be expected to be harsher with a shorter time period that the virus can maintain its viability. In addition Leung and co-workers (13) reported that up to 38% of normal breath can contain virus RNA, the presence of RNA does not equate to viable virus.

In real life it would be unrealistic to assess the amount of virus in breath or in a d-AGP. Hence, we need to look at realistic alternatives that can be readily be recorded within a primary care dental setting. It has been demonstrated (14) that 2.5micron (PM2.5), and 10 micron (PM10) particles can carry and transmit virus, particles of 2.5 microns being considered a particular problem as they can penetrate deep within the respiratory system. PM2.5 particles would expected to rapidly dehydrate and hence kill the virus. Larger PM10 could contain viable virus particles and the WHO proposed (15) 50μg/m^3^ of PM10 as this is the level considered dangerous for air pollution, not necessarily what might be necessary for transmission of a viral infection.

Hence, with little data available for the level of d-AGP particles in primary care a real-world experiment design was undertaken. The aims were two-fold. First to measure the amount of PM2.5 and PM10 aerosol particles created when creating d-AGPs (air rotor, and 3-in-1 air/water syringe). The second aim was to record the length of time required for the PM2.5 and PM10 to return to pre d-AGP levels

## Methods

The study to be undertaken in a single primary care dental surgery (Gargrave Road Dental Practice, Skipton, UK) on a volunteer patient recording PM2.5 and PM10 using a Huma-I Advanced Portable Air Quality Monitor (HI-150,)particle meter (Humatech, Bokhap-Munhwa Center, Incheon, Korea and a LifeBasis Air Quality Monitor (LifeBasis)

The first particle meter (LifeBasis) was located 10cm from the supine patient’s mouth above the patients chest and level with the oral cavity. The second particle (Huma-I) meter was situated on the side unit 1.1m from the ground 1.5 meter from the patient’s head on the opposite side of the operator. Reading for both meters for PM2.5 and PM10 were recorded at 30 second intervals. The difference between the background reading and the readings during each d-AGP was calculated.

Three d-AGPs were undertaken as follows:

1. **Air-rotor**: A Kavo super torque high speed air rota with 3 water cooling jets operated continuously for 5 minutes with a 1mm diameter fissure bur, maximum water cooling next to the lingual surface of the lower left canine with High Volume Suction (HVS) (Durr Dental, Bietigheim-Bissinggen, Germany) which was tested prior to the experiment aspirating 2.9 litres/min
2. **Air-rotor**: As above but using low volume ‘saliva ejector’ suction, No HVS
3. **3-in-1water/air syringe**: Operated into the mouth continuously for 30 seconds Tests 1 and 2 were repeated 3 times and Test 3 twice for each of the following clinic situations:
  - All doors and windows close (static air);
  - Doors closed, windows open and air conditioning fan operating (dynamic air)

## Results

The first Test performed utilised a high-volume ejector, that ejected liquid at an average speed of 2.90L/min, in well-ventilated conditions. The results for this are presented in Figure 1.

**Figure 1.**
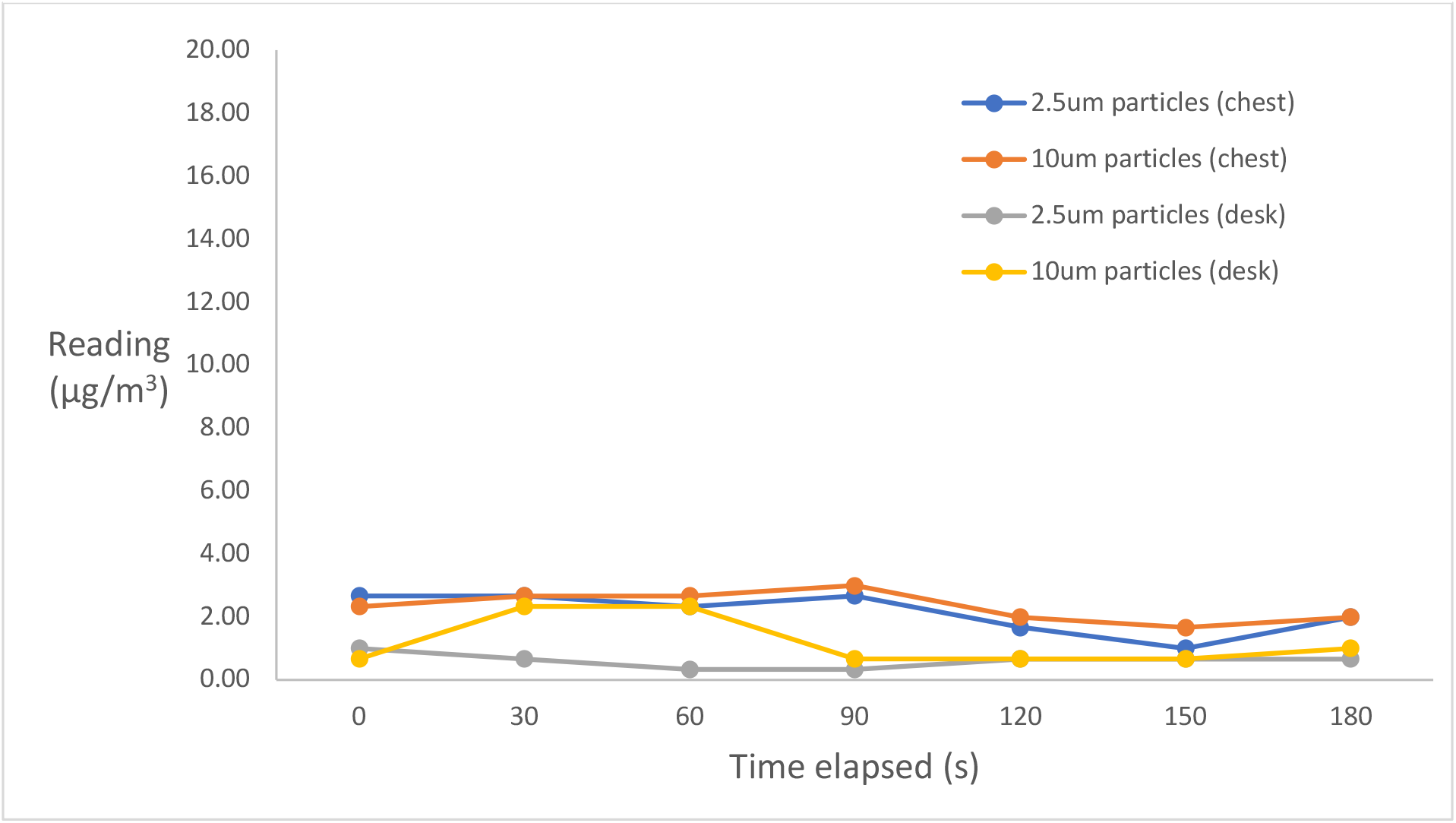
The change in measurement of concentration of particular particle sizes against time since the procedure end in a well-ventilated room, using the high-volume ejector.

**Figure 2.**
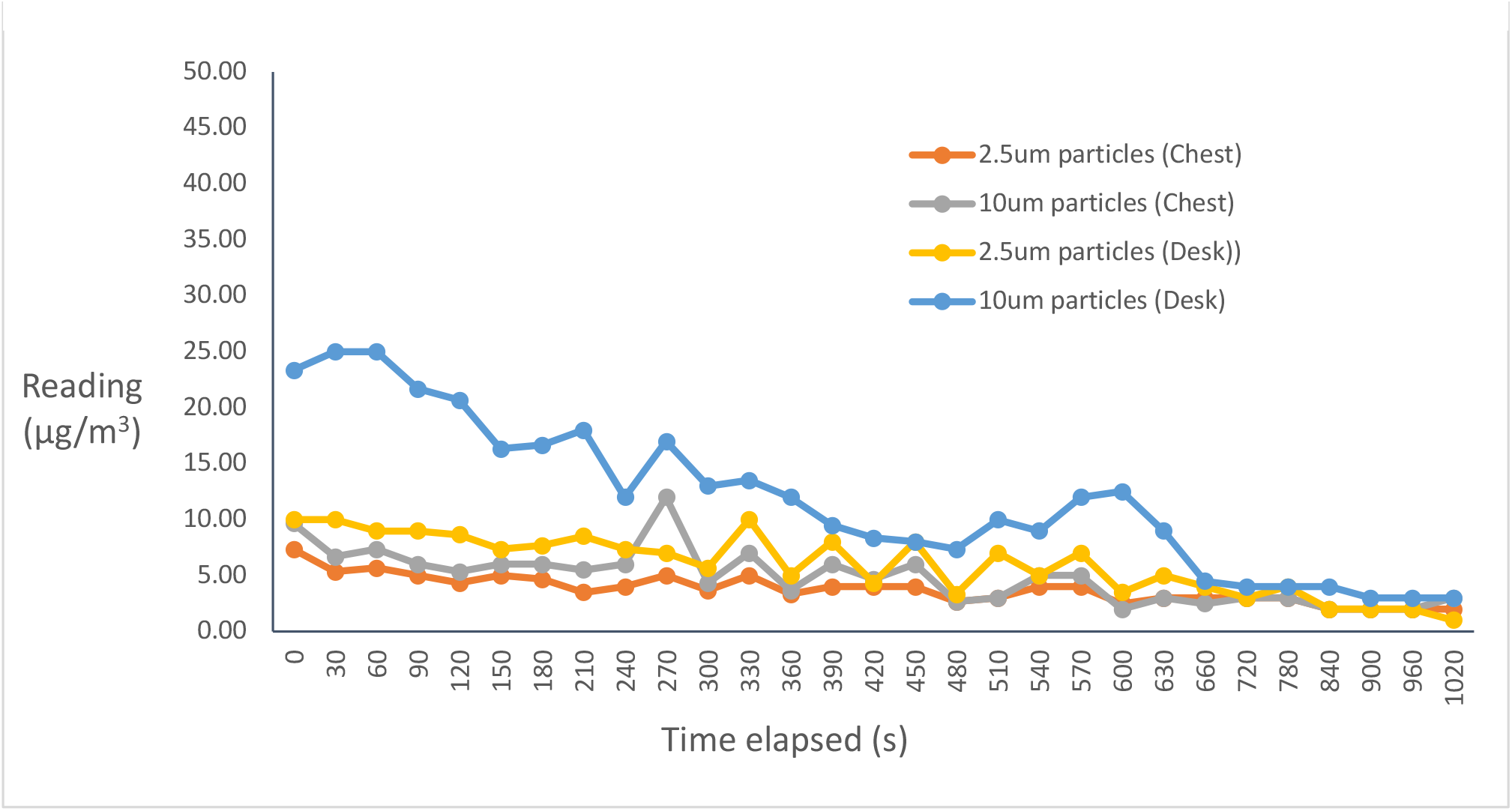
High volume suction replaced with a saliva ejector

It can be seen from the graph, that the particle readings are always well below dangerous levels. The readings wavered very little from the background concentration (time 0) for both PM2.5 and PM10.

The results from Test 2 are shown below (Figure 3).

**Figure 3.**
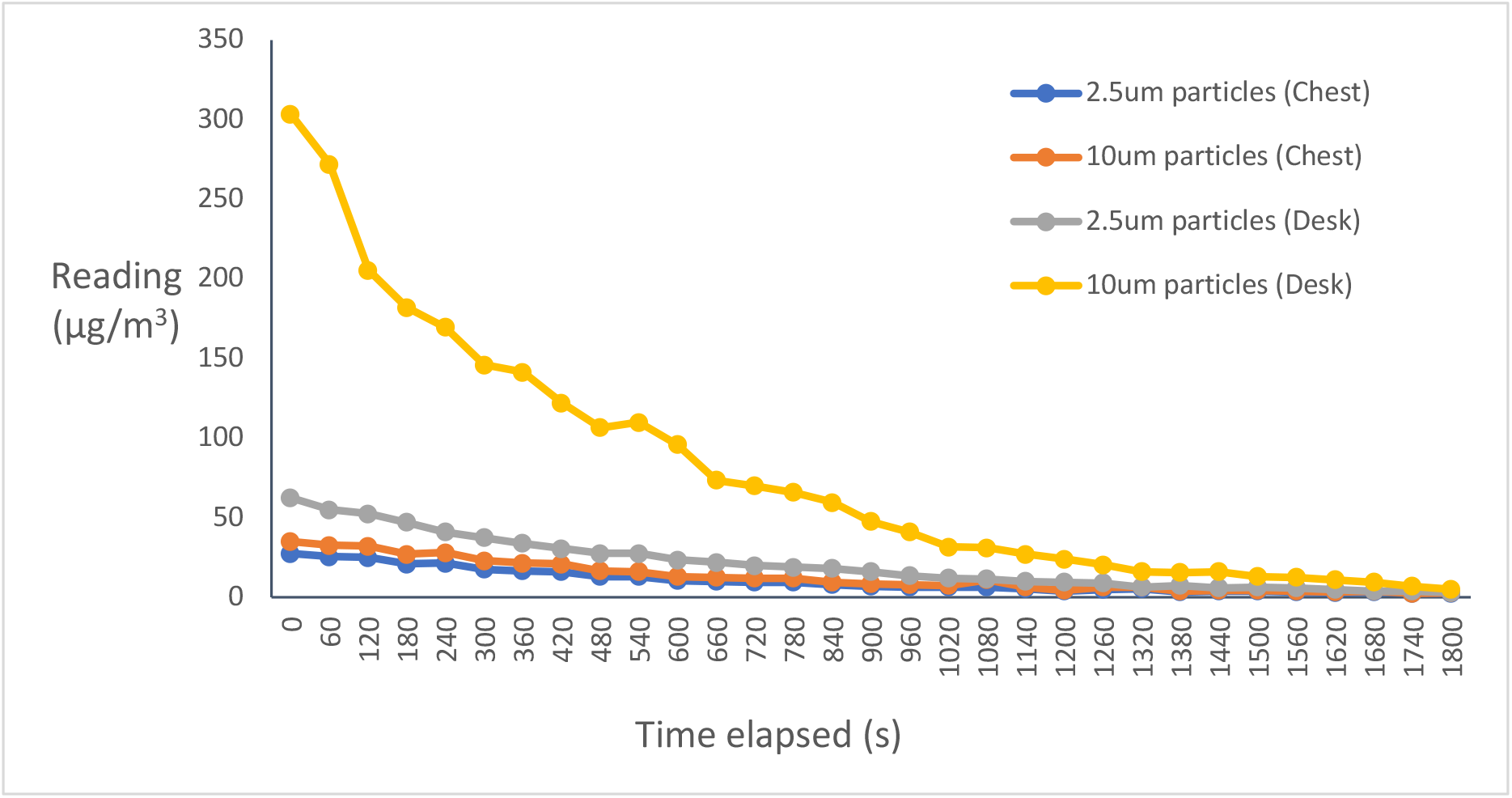
The change in measurement of concentration of particular particle sizes against the time since the procedure end in a well-ventilated room, firing the air rota into the air.

**Figure 4.**
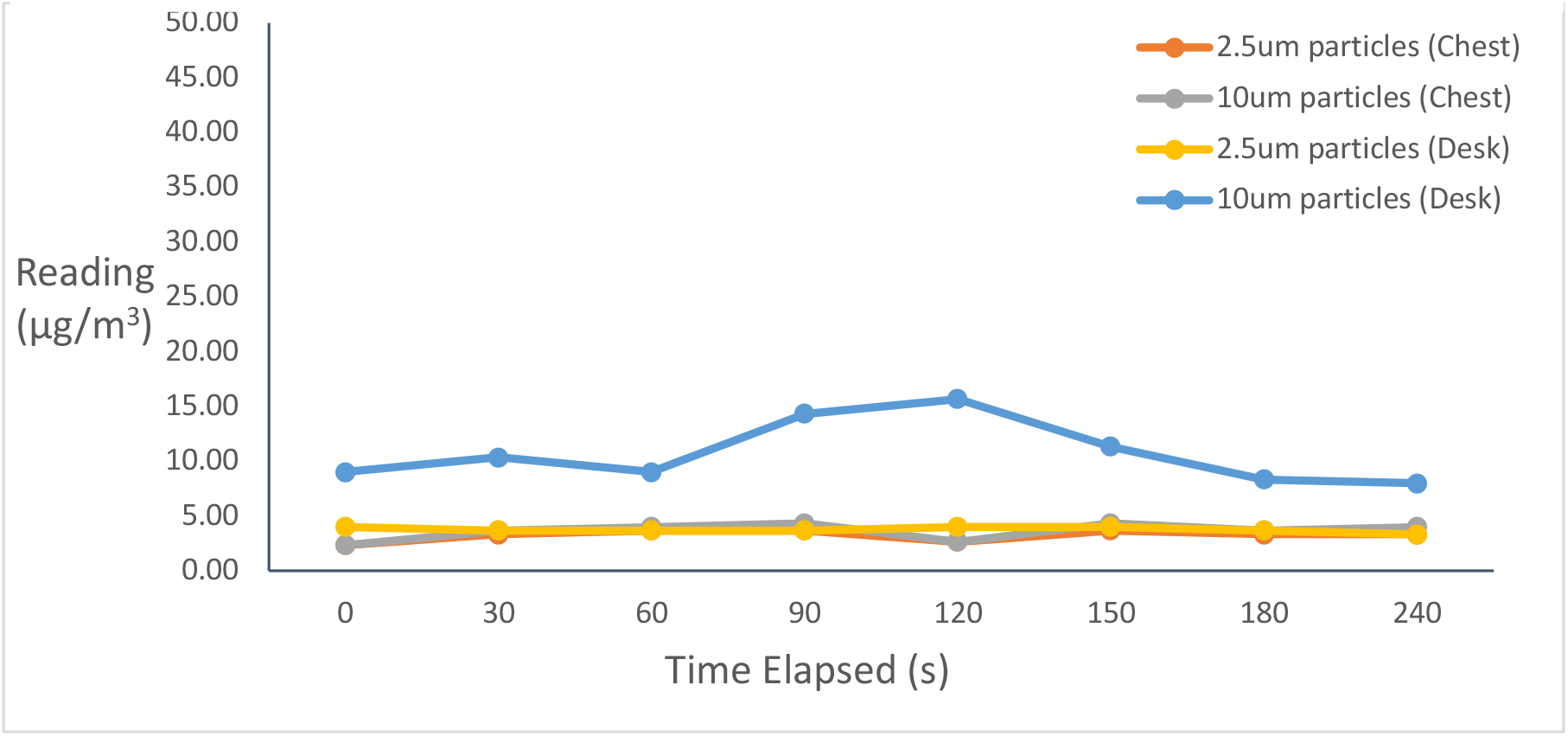
The change in measurement of concentration of particular particle sizes against time since the procedure end, without ventilation, using a high-volume ejector.

**Figure 5.**
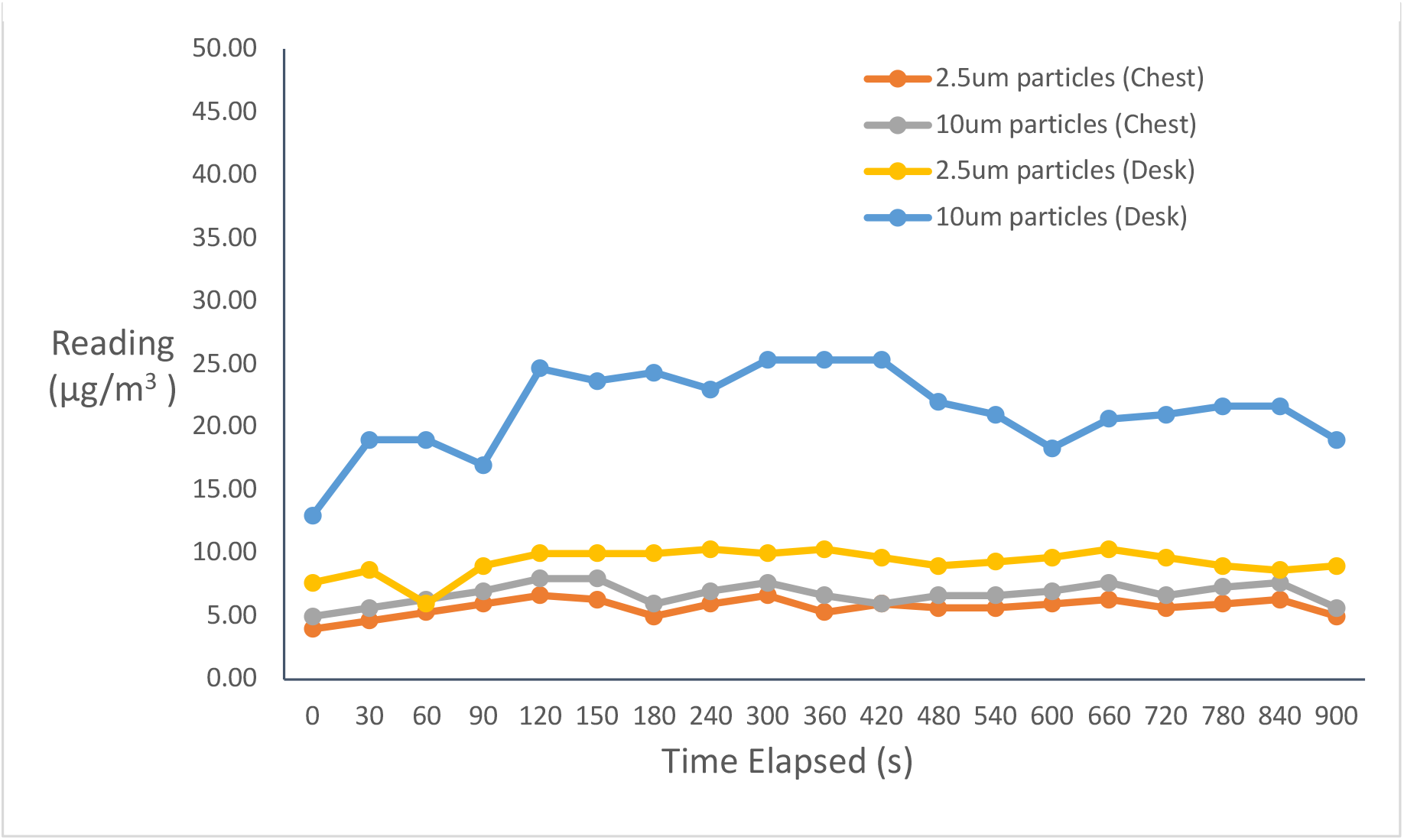
The change in measurement of concentration of particular particle sizes against the time since the procedure end, without ventilation, using a saliva ejector.

**Figure 6.**
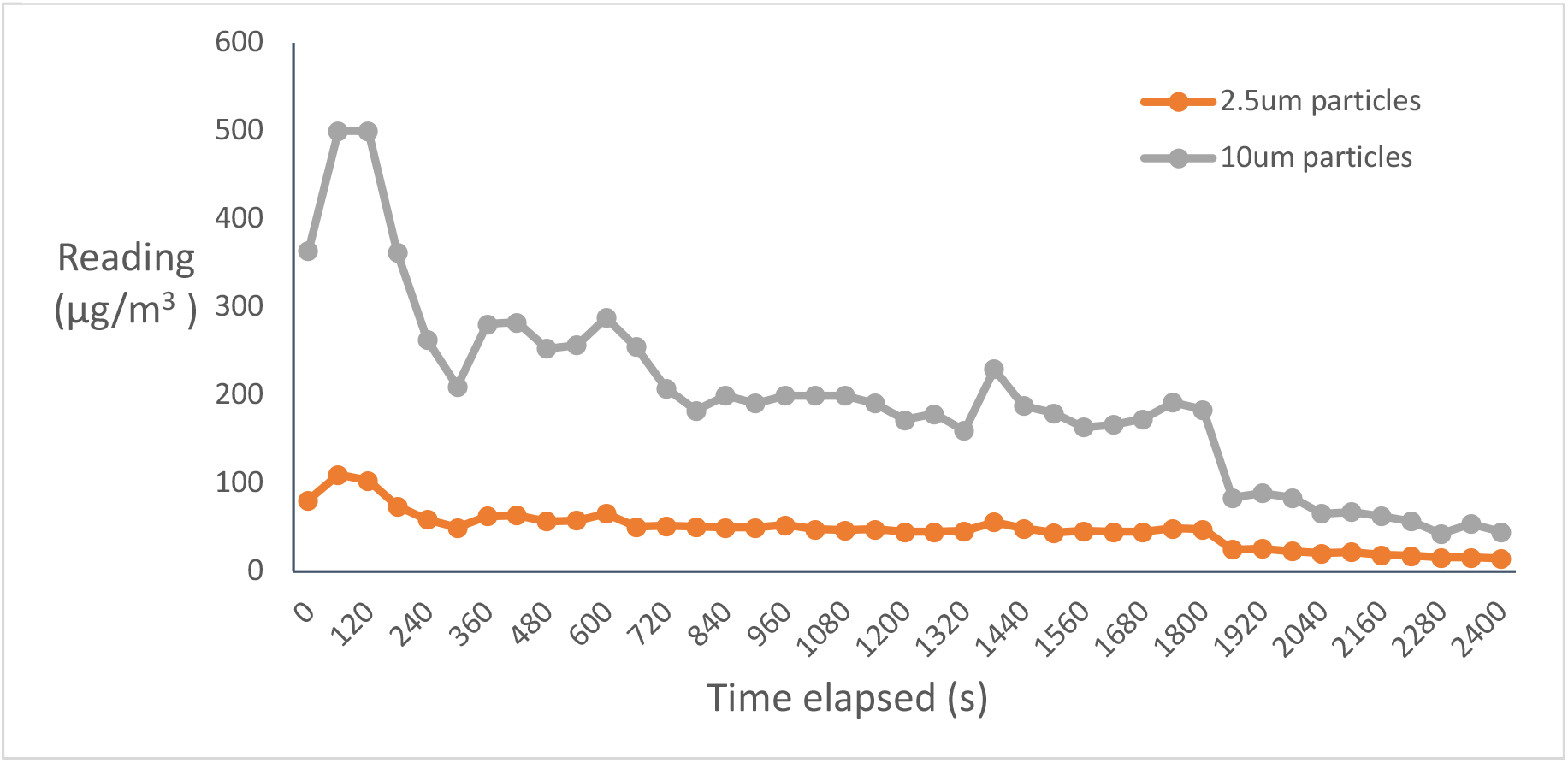
The change in measurement of concentration of particular particle sizes against the time since the procedure end, without ventilation, using a saliva ejector.

Test 2 used a saliva ejector and no HVS. The higher aerosol level also increased the time required for the particle concentration to return to background level. However, even the highest levels of particle concentration, were not near 50μg/m^3^ level that is considered generally ‘dangerous’.

Test 3 showed the effect of aiming the air rota straight up into the air out of the mouth the create the maximum amount of aerosol, on the concentration of the particle sizes under investigation.

From the graph, we can see that while there is a large 10μm particle concentration at the start of recorded readings, the value quickly falls down to safe levels by 900 seconds (15 minutes), and is indistinguishable from background concentration at 30 minutes. These levels of readings would not be seen in an ordinary procedure, as the high initial values were purposely caused by spraying the air rota directly into the air, instead of a patient’s mouth. However, even under these circumstances, the values fall to a safe level much quicker than the hour suggested by current SOPs.

The next three tables and graphs follow the same methodology, however these tests are conducted in an operating environment without ventilation, to assess its effect on particle concentration levels.

## Graph 6 - The change in measurement of concentration of particular particle sizes against the time since the procedure end, without ventilation, firing the air rota into the air

From these sets of data, we can see that the least affected by the lack of ventilation is that of Test 4, utilising a high-volume suction as the readings are always close to that of the background readings. Test 5 shows that the saliva ejector is less efficient than the high-volume ejector, with higher readings, which take much longer to dissipate due to the lack of ventilation. The final test, firing the air rota into the air without ventilation was only recorded once, as there was a large disparity between that and the readings from the chest monitor. Possible reasons for this could be the chest monitor being damaged by water when firing the air rota into the air during test 3. The readings taken show that much greater quantities of particles are detected, as well as a longer time taken for readings to decrease. The drop off at 1800 seconds (30 minutes) is due to the ventilation being introduced, again proving the effectiveness of ventilation.

## Discussion

These results demonstrate the high levels of variability when collecting data from a clinical environment using a real patient as opposed to a manikin and jig. This was demonstrated when the saliva ejector was held by the patient, which lead to a gradual reduction in aerosol across the repeated tests as the patient became better at using it. This meant that some of the values were quite different. However, this does demonstrate an important point, that patients will have different reactions to operating procedures, and those “less good” patients, i.e. those that ask for more breaks, cough or splutter more, may pose a higher risk in cross-infection, as stopping and coughing elevates the levels of particles in the air and if relying on a saliva ejector alone will result in a higher level of aerosol in the surgery. The higher aerosol level also increased the time required for the particle concentration to return to background level. However, even the highest levels of particle concentration, were not near 50μg/m^3^ level that is considered generally ‘dangerous’.

As the data shows, suggestions that an hour between patients by SOPs is excessive and unnecessary. Even when the room is not being ventilated, the simulated operating procedures the aerosol levels did not reach values of 50μg/m^3^ although we accept that this level is a pollution limit, not a cross infection control one. The study also found that with ventilation, the levels of particles of interest decrease to background levels within a few minutes of ending the d-AGP, further confirming that an hour between patients is not required. When high-volume suction is used, levels of aerosol only occasionally go above 10μg/m^3^, demonstrating it’s effectiveness in keeping aerosol levels down, improving the operating environment for both patients and dentists.

Ventilation was found to be effective in fast dispersion of aerosol. Even when procedures that generated more aerosol (saliva ejector, and when the air rota was directly into the air). When no ventilation was used, higher concentrations of aerosol were detected for a longer time period before its dispersal.

## Conclusion

This real-life experiment in a primary care surgery demonstrated the effectiveness of using high volume suction in reducing the amount of dental aerosol in the surgery. It also found that ventilation has a significant positive effect in dissipating the aerosol and the times to dissipate back to normal were within minutes and far below that of the current SOPs 1 hour.

It also appears that a particle meter is a useful instrument to have within a dental surgery to measure of aerosol levels during dental treatment, their safe dispersal and assurance of both staff and patients.

## Data Availability

All the data generated in the manuscript was our own. All references made have been noted and links provided where necessary.

## References

1. Micik RE, Miller RL, Mazzarella MA, Ryge G. Studies on dental aerobiology: I. Bacterial aerosols generated during dental procedures. Journal of dental research. 1969;48(1):49–56.

2. Abel LC, Miller RL, Micik RE, Ryge G. Studies on dental aerobiology: IV. Bacterial contamination of water delivered by dental units. Journal of Dental Research. 1971;50(6):1567–9.

3. Micik RE, Miller RL, Leong AC. Studies on dental aerobiology: III. Efficacy of surgical masks in protecting dental personnel from airborne bacterial particles. Journal of Dental Research. 1971;50(3):626–30.

4. Miller RL, Micik RE, Abel C, Ryge G. Studies on dental aerobiology: II. Microbial splatter discharged from the oral cavity of dental patients. Journal of dental research. 1971;50(3):621–5.

5. Harrel SK, Molinari J. Aerosols and splatter in dentistry: a brief review of the literature and infection control implications. Journal of the American Dental Association (1939). 2004;135(4):429–37.

6. Bennett AM, Fulford MR, Walker JT, Bradshaw DJ, Martin MV, Marsh PD. Microbial aerosols in general dental practice. British dental journal. 2000;189(12):664–7.

7. OCDOE. Standard operating procedure transition to recovery 2020 [Available from: https://www.england.nhs.uk/coronavirus/publication/preparedness-letters-for-dental-care/.

8. FGDP. Implications of COVID-19 for the safe management of general dental practice A practical guide 2020 [Available from: https://www.fgdp.org.uk/implications-covid-19-safemanagement-general-dental-practice-practical-guide.

9. GOV.UK. Decontamination in primary care dental practices (HTM 01-05) 2013 [Available from: https://www.gov.uk/government/publications/decontamination-in-primarycare-dental-practices.

10. Ge Z-y, Yang L-m, Xia J-j, Fu X-h, Zhang Y-z. Possible aerosol transmission of COVID-19 and special precautions in dentistry. Journal of Zhejiang University-SCIENCE B. 2020:1-8.

11. Organization WH. Infection prevention and control of epidemic-and pandemic-prone acute respiratory infections in health care: World Health Organization; 2014.

12. van Doremalen N, Bushmaker T, Morris DH, Holbrook MG, Gamble A, Williamson BN, et al. Aerosol and surface stability of SARS-CoV-2 as compared with SARS-CoV-1. New England Journal of Medicine. 2020;382(16):1564–7.

13. Leung NH, Chu DK, Shiu EY, Chan K-H, McDevitt JJ, Hau BJ, et al. Respiratory virus shedding in exhaled breath and efficacy of face masks. Nature medicine. 2020;26(5):676–80.

14. Coulthard P. Dentistry and coronavirus (COVID-19)-moral decision-making. British Dental Journal. 2020;228(7):503–5.

15. WHO. Ambient (outdoor) air pollution 2018 [Available from: https://www.who.int/news-room/fact-sheets/detail/ambient-(outdoor)-air-quality-and-health.

